## Supplementary Methods for "Launching a saliva-based SARS-CoV-2 surveillance testing program on a university campus"

**Supplementary methods 1**: Detailed materials and methods for saliva collection sites

To provide a saliva test specimen, the participant entered the saliva collection area, where they were directed to an available kiosk staffed by a "saliva coach," who advised on the process. Advice focused first on safety and next on how to generate an optimal specimen. Each kiosk consisted of two three-foot-wide tables, either a clear shower curtain held up by a custom-built structure or a floor-standing sneeze shield (BBASSS01, Best of Signs), an automatic no-touch hand sanitizer dispenser, and a biohazardous waste bin. First, the participant was instructed to set the kit down on the table and sanitize their hands. Next, the participant opened the kit and removed the OMNIgene (OM-505, DNA Genotek) saliva collection tube, featuring a funnel at the top. They were instructed to remove their mask and spit in the tube until the liquid portion of their saliva reached the solid black line, denoting 1mL, on the tube. Any bubbles or foam were required to be above the solid black line. The time required to produce sufficient saliva was highly variable among participants, ranging from under 3 minutes to over 10 minutes in rare cases. The participant replaced their mask, and kiosk workers observed the saliva sample to check for visible food particles, excessive color (thought to be due to substances such as coffee), excessive mucus, or excessive volume (**Appendix 1**). If a sample had too much saliva, participants were brought back to the check-in desk to receive a new collection kit and repeat the process. If a sample had excessive contaminants, participants were asked to return later that day or the following day to provide another sample.

After this quality control step, participants were instructed to close the funnel lid, which popped a bubble containing a solution proprietary to the kit manufacturer, DNAGenotek. Participants were asked to hold the tube upright and watch as the solution completely drained from the lid into the collection tube. Next, participants were instructed to unscrew the funnel top, dispose of it in a biohazardous waste container, and recap with the supplied cap. Ensuring the cap was secure, participants then inverted the tube ten times to mix the solution with the saliva. Finally, participants were directed to collect more hand sanitizer and sanitize the tube and their hands simultaneously.

At the intake station, participants scanned their tube into a Salesforce-platform laboratory information management system built by Thirdwave Analytics. Again, participants confirmed their name and date of birth. For the first eleven weeks of the study, participants dropped the tube into an open biohazard bag, held open with a stand (H13193-1000, SP Bel-Art) for no-contact sample drop-off. This bag was closed by a testing site worker and moved to a tertiary container. In the last six weeks of the study, participants were instead instructed to place the tube into a rack (93.852.200, Sarstedt Inc.), as keeping samples upright was easier for accessioning in the testing laboratory. A testing site worker wearing a gown, face shield, mask, and gloves then moved the tube into a secondary rack. This change was made to facilitate more efficient processing in the testing lab. From here, the samples were brought back to the IGI Diagnostics Lab, where they underwent the procedures outlined in Hamilton et al.

**Supplementary methods 2**: IGI FAST Exit survey text freely available at <https://docs.google.com/document/d/1-_fS3ikx6-aSAB1wqM2WYwKz_8EkZ22U0MOmPoM6aMc/edit?usp=sharing>
