## Appendices for "Launching a saliva-based SARS-CoV-2 surveillance testing program on a university campus"

**Appendix 1**: Visual inspection guide for on-site saliva sample screening. See <https://innovativegenomics.org/wp-content/uploads/2021/01/visual-inspection-guide.pdf> for the full-quality version

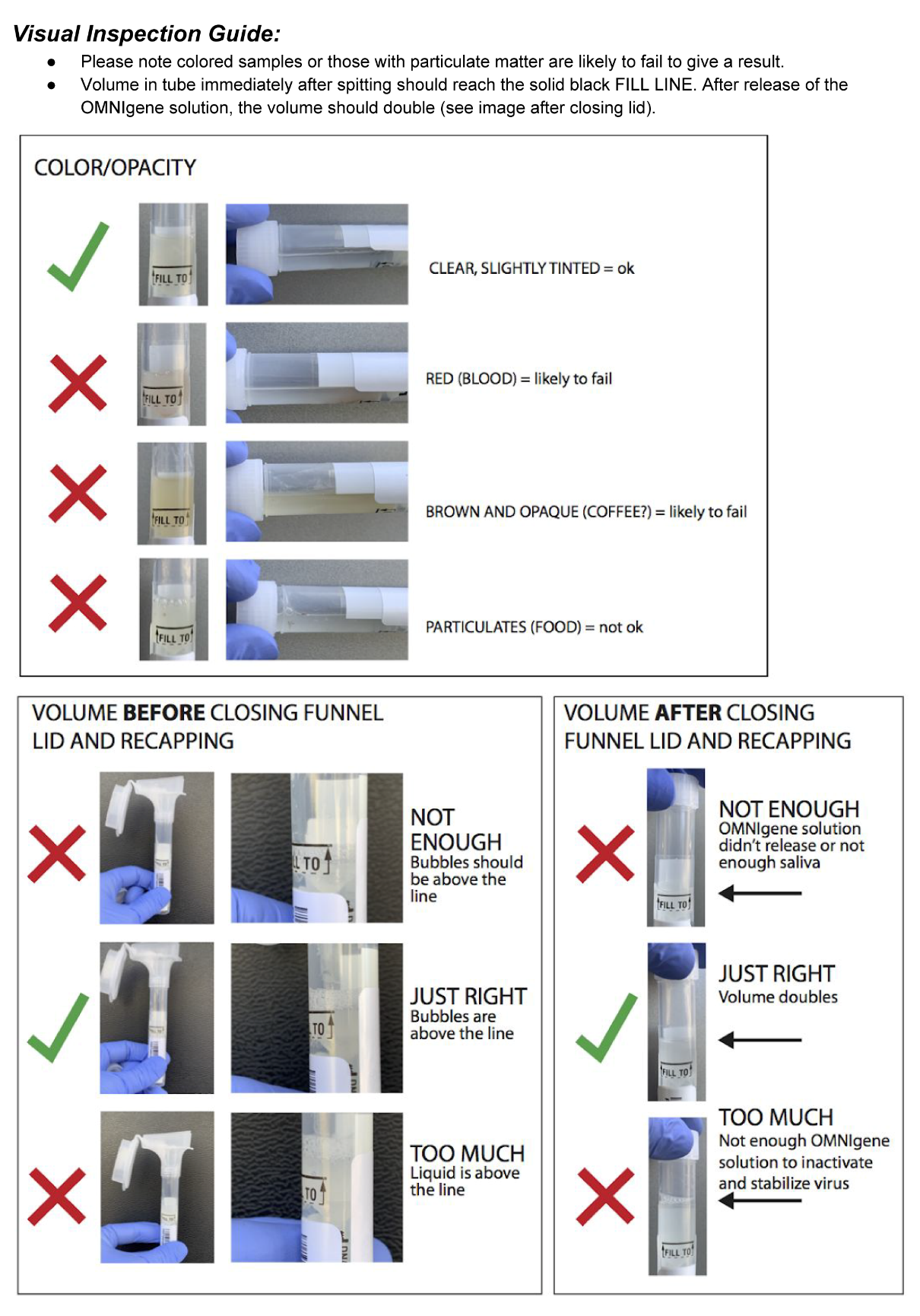

**Appendix 2:** Generic versions of the results emails used for IGI FAST

POSITIVE RESULT

*Note: Participants received a phone call from the clinician as well.*

--

Dear [NAME],

**The sample you submitted this week for the IGI FAST Study returned with a positive result for SARS-CoV-2. This means that even though you are asymptomatic, you might have COVID-19. This test was for research, and has not been FDA-approved for official medical results. These research results will not be entered into your electronic health record and do not constitute a diagnosis of COVID-19.**

I am advising that you should self-quarantine (<https://www.cdc.gov/coronavirus/2019-ncov/if-you-are-sick/quarantine-isolation.html>) and call UHS to be tested using a clinically-approved respiratory swab

Confirmation by respiratory swab is important at this stage because the sensitivity and specificity of saliva-based diagnostic tests are unknown. While there is an FDA-authorized test for saliva to be used for diagnostics elsewhere, the IGI SARS-CoV-2 Diagnostics Lab has not fully validated it using their protocol under the lab’s CLIA license. Your result will be communicated to University Health Services as a suspected COVID-19 case but will not be logged in your electronic medical record.

For students, call the UHS 24/7 phone triage at 510-643-7197 and explain that you received an inconclusive test through IGI research and they will help you take the next steps. For faculty/staff, call the UHS occupational health line at 510-332-7192 on weekdays from 10-4pm to arrange for clinical testing, letting them know you received an inconclusive research test at IGI. You are also welcomed to contact your own primary care provider to arrange for testing if you would prefer. For contractors or other individuals not employed by UC Berkeley, contact your primary care provider.

Again, please avoid contact with people, stay in your room as much as possible without visitation, clean & disinfect surfaces you touch, and wear a mask anytime you would be around people until testing is confirmed.

Please respond directly to this email to confirm you received and understand it.

--

***Appendix 2 (cont.)***

INCONCLUSIVE RESULT

*Note: Participants received a phone call from the clinician as well.*

--

Dear [NAME],

**The sample you submitted this week for the IGI FAST Study returned with an inconclusive result for SARS-CoV-2. This means that even though you are asymptomatic, you might have COVID-19. This test was for research, and has not been FDA-approved for official medical results. These research results will not be entered into your electronic health record and do not constitute a diagnosis of COVID-19.**

I am advising that you should self-quarantine (<https://www.cdc.gov/coronavirus/2019-ncov/if-you-are-sick/quarantine-isolation.html>) and call UHS to be tested using a clinically-approved respiratory swab.

Confirmation by respiratory swab is important at this stage because the sensitivity and specificity of saliva-based diagnostic tests are unknown. While there is an FDA-authorized test for saliva to be used for diagnostics elsewhere, the IGI SARS-CoV-2 Diagnostics Lab has not fully validated it using their protocol under the lab’s CLIA license. Your result will be communicated to University Health Services as a suspected COVID-19 case but will not be logged in your electronic medical record.

For students, call the UHS 24/7 phone triage at 510-643-7197 and explain that you received an inconclusive test through IGI research and they will help you take the next steps. For faculty/staff, call the UHS occupational health line at 510-332-7192 on weekdays from 10-4pm to arrange for clinical testing, letting them know you received an inconclusive research test at IGI. You are also welcomed to contact your own primary care provider to arrange for testing if you would prefer. For contractors or other individuals not employed by UC Berkeley, contact your primary care provider.

Again, please avoid contact with people, stay in your room as much as possible without visitation, clean & disinfect surfaces you touch, and wear a mask anytime you would be around people until testing is confirmed.

Please respond directly to this email to confirm you received and understand it.

--

***Appendix 2 (cont.)***

NEGATIVE RESULT

--

Dear IGI FAST Study Participant,

The sample you submitted this week for the IGI FAST Study returned with a negative result for SARS-CoV-2. This test was for research and has not been FDA-approved for official medical results. These research results will not be entered into your electronic health record.

There is an FDA-authorized test for saliva to be used for diagnostics elsewhere, however the IGI SARS-CoV-2 Diagnostics Lab has not fully validated it using their protocol under the lab’s CLIA license. While the virus that causes COVID-19, SARS-CoV-2, was not found in your sample for this week, we cannot guarantee that you do not have an active SARS-CoV-2 infection.

Continue to monitor yourself for symptoms, complete the campus [symptom screener](https://calberkeley.ca1.qualtrics.com/jfe/form/SV_3xTgcs162K19qRv) before coming to campus, maintain social distancing, and wear a mask or face covering any time you are outside or on campus (including inside campus buildings). Look for an email message regarding your next FAST study appointment.

If you have any health concerns related to COVID-19, please see <https://uhs.berkeley.edu/coronavirus-covid-19-information>

--

***Appendix 2 (cont.)***

SPECIMEN INSUFFICIENT RESULT

--

Dear IGI FAST Study Participant,

When samples, like the one you submitted this week for the IGI FAST Study, are run in the lab, they occasionally return with an “Invalid” or “Specimen Insufficient” note. Unfortunately, your sample this week returned with one of these notes. This can be due to any number of reasons. Most commonly, it is due to collection issues. Please see the recommendations below and at igi-fast.berkeley.edu/about for tips.

I’m sure that it is disappointing to not get a result from this week’s sample so we would like to offer you the opportunity to come back in before your next appointment in two weeks. Please feel free to drop-in at the same collection site any time [DATES AND TIMES AVAILABLE OVER THE NEXT WEEK]. You do not need an appointment. Just let the check-in personnel know that you are there for retesting. Otherwise, you are welcome to just wait to schedule your next appointment through our normal system for roughly two weeks from now.

Please note that this note is different from an inconclusive result. While the note here indicates that there was an issue with the sample, an inconclusive result indicates that viral RNA was successfully extracted and some signal associated with SARS-CoV-2 was detected but did not reach the threshold necessary to label the sample as positive. This is why an inconclusive result necessitates confirmatory clinical testing while the result here does not.

To maximize the chances of successful sampling, we have the following general recommendations for saliva collection:

- Make sure to hydrate well during the day, but not eat, drink (including water), smoke, or vape for at least 30 minutes prior to your appointment. We have noticed that consuming plain water 30-60 minutes prior to your appointment helps to rinse out any residual food or drink like coffee. At this point, we do **not** recommend brushing your teeth or using mouth wash within this time frame.
- Try to conger saliva from the front of your mouth instead of the back. Do not attempt to cough up mucus or sputum to add to the volume in your sample. The viscosity of mucus interferes with the protocols in the lab.
- Make sure to fill your tube up to, but not exceeding the line. There shouldn’t be any bubbles below the line. A kiosk volunteer is happy to check the levels you have if you are unsure.
- Make sure to fully close the funnel cap until you hear a pop or click, which indicates that a preservative fluid has been released into your sample. Keeping the tube upright, watch to make sure all the fluid runs down from the cap into the tube. Kiosk personnel can help confirm whether or not this has happened.
- Take as much time as you need at the kiosk. Saliva production rates can vary throughout the day and by person-to-person. You do not need to feel rushed at any point. If you need any help, please let any of the kiosk personnel know.

If you have any health concerns related to COVID-19, please see <https://uhs.berkeley.edu/coronavirus-covid-19-information>

**Appendix 3:** Instructional sheet (front and back) for at-home saliva collection. See <https://innovativegenomics.org/wp-content/uploads/2021/01/At-home-saliva-v7-RGB-4-x-9-Front-and-back.pdf> for full quality version.

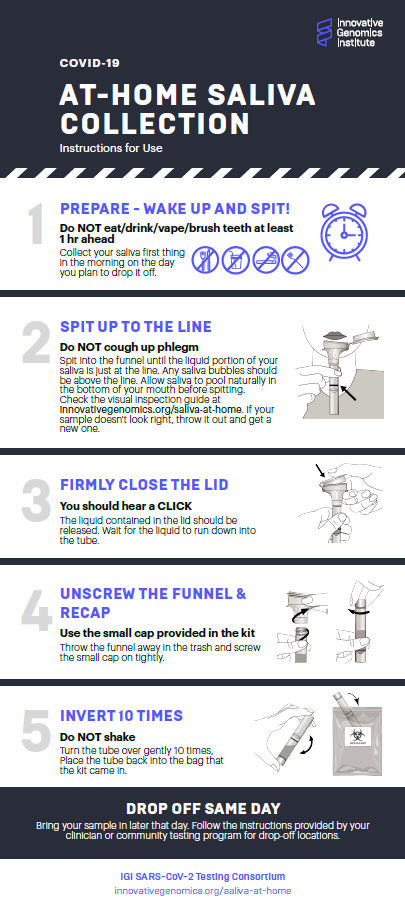

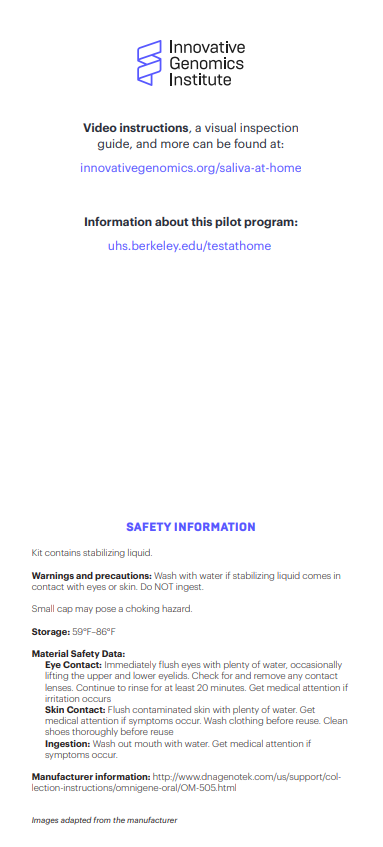
