## Supplementary Data for "Launching a saliva-based SARS-CoV-2 surveillance testing program on a university campus"

**Supplementary data 1**: Daily IGI FAST results and estimates of asymptomatic and presymptomatic SARS-CoV-2 infection prevalence in the City of Berkeley, derived from the ‘covidestim’ R package.

|  | **IGI FAST Data** | | | | |  | **Estimates of community prevalence** |
| --- | --- | --- | --- | --- | --- | --- | --- |
| **Collection Date** | **Negative** | **Positive** | **Inconclusive** | **Insufficient** | **Total** |  | **Asymptomatic and presymptomatic prevalence (%, 95% CI)** |
| 6/23/2020 | 106 | 0 | 1 | 57 | 164 |  | 0.14% (0.06%, 0.42%) |
| 6/25/2020 | 131 | 0 | 0 | 42 | 173 |  | 0.12% (0.05%, 0.42%) |
| 6/26/2020 | 130 | 0 | 0 | 27 | 157 |  | 0.12% (0.05%, 0.39%) |
| 6/30/2020 | 173 | 0 | 0 | 63 | 236 |  | 0.1% (0.04%, 0.32%) |
| 7/2/2020 | 81 | 0 | 0 | 52 | 133 |  | 0.09% (0.04%, 0.3%) |
| 7/6/2020 | 212 | 0 | 1 | 64 | 277 |  | 0.08% (0.03%, 0.25%) |
| 7/8/2020 | 205 | 0 | 1 | 71 | 277 |  | 0.07% (0.03%, 0.22%) |
| 7/13/2020 | 266 | 0 | 0 | 16 | 282 |  | 0.07% (0.03%, 0.2%) |
| 7/15/2020 | 214 | 0 | 0 | 9 | 223 |  | 0.06% (0.03%, 0.2%) |
| 7/20/2020 | 313 | 0 | 1 | 6 | 320 |  | 0.07% (0.03%, 0.2%) |
| 7/22/2020 | 218 | 0 | 0 | 36 | 254 |  | 0.07% (0.03%, 0.2%) |
| 7/27/2020 | 307 | 0 | 0 | 11 | 318 |  | 0.07% (0.03%, 0.24%) |
| 7/29/2020 | 223 | 1 | 0 | 10 | 234 |  | 0.08% (0.03%, 0.23%) |
| 8/3/2020 | 312 | 1 | 0 | 4 | 317 |  | 0.08% (0.03%, 0.25%) |
| 8/5/2020 | 287 | 0 | 2 | 6 | 295 |  | 0.08% (0.03%, 0.26%) |
| 8/10/2020 | 298 | 0 | 1 | 23 | 322 |  | 0.08% (0.04%, 0.26%) |
| 8/12/2020 | 267 | 0 | 1 | 19 | 287 |  | 0.08% (0.04%, 0.27%) |
| 8/17/2020 | 229 | 0 | 0 | 2 | 231 |  | 0.09% (0.04%, 0.26%) |
| 8/19/2020 | 359 | 0 | 4 | 9 | 372 |  | 0.09% (0.04%, 0.26%) |
| 8/24/2020 | 242 | 0 | 1 | 6 | 249 |  | 0.08% (0.03%, 0.26%) |
| 8/26/2020 | 387 | 1 | 2 | 63 | 453 |  | 0.08% (0.03%, 0.25%) |
| 9/21/2020 | 440 | 1 | 0 | 12 | 453 |  | 0.02% (0.01%, 0.07%) |
| 9/22/2020 | 445 | 1 | 1 | 7 | 454 |  | 0.02% (0.01%, 0.08%) |
| 9/23/2020 | 270 | 0 | 0 | 1 | 271 |  | 0.02% (0.01%, 0.08%) |
| 9/24/2020 | 181 | 0 | 0 | 13 | 194 |  | 0.02% (0.01%, 0.08%) |
| 9/25/2020 | 314 | 0 | 0 | 3 | 317 |  | 0.02% (0.01%, 0.09%) |
| 9/28/2020 | 156 | 0 | 0 | 6 | 162 |  | 0.03% (0.01%, 0.1%) |
| 9/29/2020 | 97 | 0 | 1 | 2 | 100 |  | 0.03% (0.01%, 0.11%) |
| 9/30/2020 | 118 | 0 | 0 | 3 | 121 |  | 0.03% (0.01%, 0.12%) |
| 10/1/2020 | 91 | 0 | 0 | 6 | 97 |  | 0.03% (0.01%, 0.13%) |
| 10/5/2020 | 355 | 0 | 0 | 41 | 396 |  | 0.04% (0.02%, 0.16%) |
| 10/6/2020 | 408 | 0 | 0 | 5 | 413 |  | 0.05% (0.02%, 0.18%) |
| 10/7/2020 | 235 | 0 | 0 | 11 | 246 |  | 0.05% (0.02%, 0.21%) |
| 10/8/2020 | 199 | 0 | 0 | 4 | 203 |  | 0.06% (0.02%, 0.21%) |
| 10/9/2020 | 300 | 0 | 1 | 4 | 305 |  | 0.06% (0.02%, 0.22%) |
| 10/12/2020 | 298 | 0 | 0 | 10 | 308 |  | 0.08% (0.03%, 0.29%) |
| 10/13/2020 | 198 | 0 | 0 | 5 | 203 |  | 0.08% (0.03%, 0.32%) |
| 10/14/2020 | 214 | 0 | 0 | 4 | 218 |  | 0.09% (0.03%, 0.34%) |
| 10/15/2020 | 157 | 0 | 0 | 1 | 158 |  | 0.1% (0.04%, 0.36%) |
| 10/16/2020 | 201 | 0 | 2 | 8 | 211 |  | 0.1% (0.04%, 0.37%) |
| 10/19/2020 | 230 | 0 | 0 | 0 | 230 |  | 0.12% (0.04%, 0.44%) |
| 10/20/2020 | 294 | 0 | 0 | 1 | 295 |  | 0.13% (0.05%, 0.47%) |
| 10/21/2020 | 190 | 0 | 0 | 1 | 191 |  | 0.14% (0.05%, 0.52%) |
| 10/22/2020 | 178 | 0 | 0 | 0 | 178 |  | 0.15% (0.05%, 0.55%) |
| 10/23/2020 | 185 | 0 | 0 | 2 | 187 |  | 0.16% (0.06%, 0.59%) |
| 10/26/2020 | 136 | 0 | 0 | 4 | 140 |  | 0.2% (0.07%, 0.66%) |
| 10/27/2020 | 128 | 0 | 0 | 2 | 130 |  | 0.23% (0.08%, 0.72%) |
| 10/28/2020 | 101 | 0 | 0 | 0 | 101 |  | 0.24% (0.09%, 0.76%) |
| 10/29/2020 | 105 | 0 | 1 | 9 | 115 |  | 0.25% (0.1%, 0.8%) |
